# Antibody testing for COVID-19: A report from the National COVID Scientific Advisory Panel

**DOI:** 10.1101/2020.04.15.20066407

**Authors:** National COVID Testing Scientific Advisory Panel, Emily R Adams, Mark Ainsworth, Rekha Anand, Monique I Andersson, Kathryn Auckland, J Kenneth Baillie, Eleanor Barnes, Sally Beer, John Bell, Tamsin Berry, Sagida Bibi, Miles Carroll, Senthil Chinnakannan, Elizabeth Clutterbuck, Richard J Cornall, Derrick W Crook, Thushan De Silva, Wanwisa Dejnirattisai, Kate E Dingle, Christina Dold, Alexis Espinosa, David W Eyre, Helen Farmer, Maria Fernandez Mendoza, Dominique Georgiou, Sarah J Hoosdally, Alistair Hunter, Katie Jeffrey, Paul Klenerman, Julian Knight, Clarice Knowles, Andrew J Kwok, Ullrich Leuschner, Robert Levin, Chang Liu, Cesar Lopez-Camacho, Jose Carlos Martinez Garrido, Philippa C Matthews, Hannah McGivern, Alexander J Mentzer, Jonathan Milton, Juthathip Mongkolsapaya, Shona C Moore, Marta S Oliveira, Fiona Pereira, Elena Perez Lopez, Timothy Peto, Rutger J Ploeg, Andrew Pollard, Tessa Prince, David J Roberts, Justine K Rudkin, Veronica Sanchez, Gavin R Screaton, Malcolm G Semple, Donal T Skelly, Jose Slon-Campos, Elliot Nathan Smith, Alberto Jose Sobrino Diaz, Julie Staves, David Stuart, Piyada Supasa, Tomas Surik, Hannah Thraves, Pat Tsang, Lance Turtle, A Sarah Walker, Beibei Wang, Charlotte Washington, Nicholas Watkins, James Whitehouse

**Affiliations:** Liverpool School of Tropical Medicine; Oxford University Hospitals NHS Foundation Trust; NHSBT Birmingham; Department of Microbiology, Oxford University Hospital NHS Foundation Trust; The Wellcome Centre for Human Genetics, University of Oxford; Roslin Institute, University of Edinburgh; Nuffield Department of Medicine, University of Oxford; Department of Medicine, University of Oxford; Department of Health and Social Care, University of Oxford; Oxford Vaccine group, Department of Pediatrics, University of Oxford; Nuffield Department of Medicine, Centre of Tropical Medicine and Global Health and Public Health England; Oxford Vaccine Group, Department of Paediatrics, University of Oxford; NIHR Oxford Biomedical Research Centre; Department of Infection, Immunity and Cardiovascular, Disease, The Medical School, University of Sheffield; Wellcome Centre for Human Genetics, Nuffield Department of Medicine, University of Oxford; NIHR Oxford Biomedical Research Centre, University of Oxford; Big Data Institute, University of Oxford; NHSBT Basildon; Department of Clinical Medicine, Oxford University Hospitals NHS Foundation Trusts; Nuffield Department so Medicine, University of Oxford; NHSBT Oxford; Worthing Hospital, Worthing, West Sussex; Wellcome Centre of Genetics, Nuffield Department of Medicine, University of Oxford; Nuffield Department of Medicine, University of Medicine; Nuffield Department of Surgical Sciences, University of Oxford and UK QUOD Consortium; Wellcome Centre for Human Genetics, University of Oxford; NIHR Health Protection Research Unit for Emerging and Zoonotic Infections, University of Liverpool; Imperial College London; NIHR Oxford Biomedical Research centre, University of Oxford; Department of Paediatrics, University of Oxford; Nuffield Department of Population Health & Big Data Institute, University of Oxford; Health Protection Unit In Emerging and Zoonotic Infection, University of Liverpool; Nuffield Department of Clinical Neurosciences, University of Oxford; University of Oxford; Oxford University Hospitals; Wellcome Centre for Human Genetics, Nuffield Department of Medicine; NHSBT, Birmingham; NHSBT, Cambridge

**Keywords:** COVID-19, SARS-CoV-2, serology, IgG, IgM, antibodies, immunoassay, ELISA, lateral flow, exposure, epidemiology

## Abstract

**Background:** The COVID-19 pandemic caused >1 million infections during January-March 2020. There is an urgent need for reliable antibody detection approaches to support diagnosis, vaccine development, safe release of individuals from quarantine, and population lock-down exit strategies. We set out to evaluate the performance of ELISA and lateral flow immunoassay (LFIA) devices.

**Methods:** We tested plasma for COVID (SARS-CoV-2) IgM and IgG antibodies by ELISA and using nine different LFIA devices. We used a panel of plasma samples from individuals who have had confirmed COVID infection based on a PCR result (n=40), and pre-pandemic negative control samples banked in the UK prior to December-2019 (n=142).

**Results:** ELISA detected IgM or IgG in 34/40 individuals with a confirmed history of COVID infection (sensitivity 85%, 95%CI 70-94%), vs. 0/50 pre-pandemic controls (specificity 100% [95%CI 93-100%]). IgG levels were detected in 31/31 COVID-positive individuals tested ≥10 days after symptom onset (sensitivity 100%, 95%CI 89-100%). IgG titres rose during the 3 weeks post symptom onset and began to fall by 8 weeks, but remained above the detection threshold. Point estimates for the sensitivity of LFIA devices ranged from 55-70% versus RT-PCR and 65-85% versus ELISA, with specificity 95-100% and 93-100% respectively. Within the limits of the study size, the performance of most LFIA devices was similar.

**Conclusions:** Currently available commercial LFIA devices do not perform sufficiently well for individual patient applications. However, ELISA can be calibrated to be specific for detecting and quantifying SARS-CoV-2 IgM and IgG and is highly sensitive for IgG from 10 days following first symptoms.

## INTRODUCTION

The first cases of infection with a novel coronavirus (SARS-CoV-2) causing Coronavirus Infectious Disease (COVID) emerged in Wuhan, China on December 31st, 2019 [1]. Despite intensive containment efforts, there was rapid international spread and three months later, there had been over 1 million confirmed infections and 60,000 reported deaths [2]. Containment efforts have relied heavily on population quarantine (‘ lock-down’) measures to restrict movement and reduce individual contacts [3,4]. To develop public health strategies for exit from lock-down, diagnostic testing urgently needs to be scaled-up, including both mass screening and screening of specific high-risk groups (contacts of confirmed cases, and healthcare workers and their families), in parallel with collecting data on recent and past infection at individual and population levels [2].

Laboratory diagnosis of infection has mostly been based on real-time RT-PCR, typically targeting the viral RNA-dependent RNA polymerase (RdRp) or nucleocapsid (N) genes using swabs collected from the upper respiratory tract [5,6]. This requires specialist equipment, skilled laboratory staff and PCR reagents, creating diagnostic delays. RT-PCR from upper respiratory tract swabs may also be falsely negative due to quality or timing; viral loads in upper respiratory tract secretions peak in the first week of symptoms [7], but may have declined below the limit of detection in those presenting later [8]. In individuals who have recovered, RT-PCR provides no information about prior exposure or immunity.

In contrast, assays that reliably detect antibody responses specific to SARS-CoV-2 could contribute to diagnosis of acute infection (via rises in IgM and IgG levels) and to identifying those infected with or without symptoms and recovered (via persisting IgG) [9]. Receptor-mediated viral entry to host cells occurs through interactions between the unique and highly-conserved viral spike (S) glycoprotein and the ACE2 cell receptor [10]. This S protein is the primary target of specific neutralising antibodies, and current SARS-CoV-2 serology assays therefore typically seek to identify these antibodies (Figure 1A-C). Rapid lateral flow immunoassay (LFIA) devices provide a quick, point-of-care approach to antibody testing. A sensitive and specific antibody assay could directly contribute to early identification and isolation of cases, address unknowns regarding the extent of infection to inform mathematical models and support individual or population-level release from lock-down. Laboratory-based ELISA platforms have also been evaluated as an approach to detection and quantification of SARS-CoV-antibodies [11].

However, before either laboratory assays or LFIA devices can be widely deployed, their performance needs to be carefully evaluated (Figure 1D,E) [12]. We therefore compared a novel laboratory-based ELISA assay with nine commercially-available LFIA devices using samples from patients with RT-PCR-confirmed infection, and negative pre-pandemic samples.

## METHODS

### Research Reporting

We have provided a STARD checklist (essential items for reporting diagnostic accuracy studies); Table S1.

### Samples

142 plasma samples designated seronegative for SARS-CoV-2 were collected from adults (≥18 years) in the UK before December 2019 (Table S2, including demographic details) from three ethically approved sources: healthy blood donors, organ donors on ICU following cerebral injury and healthy volunteers from a vaccine study.

Forty plasma samples were collected from adults positive for SARS-CoV-2 by RT-PCR from an upper respiratory tract (nose/throat) swab tested in accredited laboratories (Table S2). Acute (≤28 days from symptom onset) and convalescent samples (>28 days) were included to optimise detection of SARS-CoV-2 specific IgM and IgG respectively (Figure 1B). Acute samples were collected from patients a median 10 (range 4-27) days from symptom onset (n=16), and from recovering healthcare workers median 13 [range 8-19] days after first symptoms; (n=6). Convalescent samples were collected from adults a median 48 [range 31-62] days after symptom onset and/or date of positive throat swab (n=18). Further sample details are provided in Supplementary Material.

Cases were classified following WHO criteria as critical (respiratory failure, septic shock, and/or multiple organ dysfunction/failure); severe (dyspnoea, respiratory frequency ≥30/minute, blood oxygen saturation ≤93%, PaO2/FiO2 ratio <300, and/or lung infiltrates >50% of the lung fields within 24-48 hours); or otherwise mild [13]. Among 22 acute cases, 9 were critical, 4 severe and 9 mild. All but one convalescent individual had mild disease; the other was asymptomatic and screened during enhanced contact tracing.

### Enzyme-linked immunosorbent assay (ELISA)

We developed a novel ELISA targeting the SARS-CoV-2 spike protein. Recombinant SARS-CoV-2 trimeric spike protein was constructed as described [14], using mammalian codon optimized SARS2 Spike (1-1208, Genbank no. MN908947) with a GSAS substitution at the furin cleavage site (aa 682-685) and double proline substitution at aa 986-987. The C-terminal was followed by T4 fibritin motif, an HRV3C protease cleavage site, a TwinStrep Tag and an 8-HisTag. The gene was cloned into a pHLsec and expressed in 293T cells. The HIS trap HP column (cat no 17524701; Cytiva) was used to purify the recombinant S protein.

We used ELISA to detect antibodies to the S protein. MAXISORP immunoplates (442404; NUNC) were coated with StrepMAB-Classic (2-1507-001;iba). Plates were blocked with 2% skimmed milk in PBS for one hour and then incubated with 0.125ug of soluble trimeric SARS-CoV-2 trimeric S protein or 2% skimmed milk in phosphate buffered saline. After one hour, plasma was added at 1:50 dilution, followed by ALP-conjugated anti-human IgG (A9544; Sigma) at 1:10,000 dilution or ALP-conjugated anti-human IgM (A9794; Sigma) at 1:5,000 dilution. The reaction was developed by the addition of PNPP substrate and stopped with 1.0 M NaOH. The absorbance was measured at 405nm after 90 minutes, and a final optical density (OD) value was calculated by subtracting the background (skimmed milk) from the test value. The ELISA assay takes 5-6 hours to perform with an experienced operator being able to process up to five 96-well plates (480 samples including relevant controls).

### Lateral flow immunoassays (LFIA)

We tested LFIA devices designed to detect IgM, IgG or total antibodies to SARS-CoV-2 produced by nine manufacturers short-listed as a testing priority by the UK Government Department of Health and Social Care (DHSC), based on appraisals of device provenance and available performance data. Individual manufacturers did not approve release of device-level data, so device names are anonymised.

Testing was performed in strict accordance with the manufacturer’ s instructions for each device. Typically, this involved adding 5-20µl of plasma to the sample well, and 80-100µl of manufacturer’ s buffer to an adjacent well, followed by incubation at room temperature for 10-15 minutes. The result was based on the appearance of coloured bands, designated as positive (control and test bands present), negative (control band only), or invalid (no band, absent control band, or band in the wrong place) (Figure 1C).

We recorded results in real-time on a password-protected electronic database, using pseudonymised sample identifiers, capturing the read-out from the device (positive/negative/invalid), operator, device, device batch number, and a timestamped photograph of the device.

### Testing protocol

We tested 90 samples using ELISA to quantify IgM and IgG antibody in plasma designated SARS-CoV-2 negative (n=50) and positive (n=40). All positive samples were included and an unstratified random sample of negative plasma from healthy blood donors (n=23) and organ donors (n=27). We tested the nine different LFIA devices using between 39-165 individual plasma samples (8-23 and 31-142 samples designated SARS-CoV-2 positive and negative, respectively, Table S3). Total numbers varied according to the number of devices supplied to the DHSC; samples were otherwise selected at random.

### Statistical analysis

Analyses were conducted using R (version 3.6.3) and Stata (version 15.1), with additional plots generated using GraphPad Prism (version 8.3.1). Binomial 95% confidence intervals (CI) were calculated for all proportions. The association between ELISA results and time since symptom onset, severity, need for hospital admission and age was estimated using multivariable linear regression, without variable selection. Non-linearity in relationships with continuous factors was included via natural cubic splines. Differences between LFIA devices were estimated using mixed effects logistic regression models, allowing for each device being tested on overlapping sample sets. Differences between devices were compared with Benjamini-Hochberg corrected p-value thresholds. (Further details in Supplementary Material.)

### Ethical approval and role of the funding source

Our work was undertaken with ethical approval from the National Health Service Blood and Transplant (NHSBT) ethics, providing donor consent for plasma use; NIHR Biobank REC agreement (REC 13/NW/0017; IRAS 87824); International Severe Acute Respiratory and Emerging Infection Consortium (‘ ISARIC’) approval by the South Central (Oxford C) Research Ethics Committee in England (Ref: 13/SC/0149), and Scotland A Research Ethics Committee in Scotland (Ref: 20/SS/0028). The UK Government DHSC selected the lateral flow devices for testing as described above. Otherwise, the funders had no role in study design or in the collection, analysis, and interpretation of data. Authors from DHSC contributed to writing of the report and in the decision to submit the paper for publication.

## RESULTS

### Detection of SARS-CoV-2 IgM and IgG antibody by ELISA

Forty positive (RT-PCR-confirmed SARS-CoV-2 infection) and 50 designated negative (pre-pandemic) plasma samples were tested by ELISA to characterise antibody profiles. Negative samples had median optical density (OD) for IgM of −0.0001 (arbitrary units) (range −0.14 to 0.06) and for IgG −0.01 (range −0.38 to 0.26). The median IgM reading in 40 positive samples was 0.18 (range −0.008 to 1.13; Kruskal-Wallis p<0.001 vs. negative) and IgG median 3.0 (range −0.2 to 3.5; p<0.001).

As safe individual release from lock-down is a major application for serological testing, we chose OD thresholds that maintained 100% specificity (95%CI 93-100%), while maximising sensitivity. Using thresholds of 0.07 for IgM and 0.4 for IgG (3 and 5 standard deviations above the negative mean respectively; Figure 2A,B), the IgG assay had 85% sensitivity (95%CI 70-94%; 34/40) vs. RT-PCR diagnosis. All six false-negatives were from samples taken within 9 days of symptom onset (Figure 2D). IgG levels were detected in 31/31 RT-PCR-positive individuals tested ≥10 days after symptom onset (sensitivity 100%, 95%CI 89-100%). The IgM assay sensitivity was lower at 70% (95%CI 53-83%; 28/40). All IgG false-negatives were IgM-negative. No patient was IgM-positive and IgG-negative.

Considering the relationship between IgM and IgG titres and time since symptom onset (Figures 2C,D), univariable regression models showed IgG antibody titres rising over the first 3 weeks from symptom onset. The lower bound of the pointwise 95%CI for the mean expected titre crosses our OD threshold between days 6-7 (Figure 2D). However, given sampling variation, test performance is likely to be optimal from several days later. IgG titres fell during the second month after symptom onset but remained above the OD threshold. No temporal association was observed between IgM titres and time since symptom onset (Figure 2C). There was no evidence that SARS-2-CoV severity, need for hospital admission or patient age were associated with IgG or IgM titres in multivariable models (p>0.1, Table S4).

### Detection of SARS-CoV-2 antibodies by LFIA vs. RT-PCR

We first considered performance of the nine different LFIA devices using RT-PCR-confirmed cases as the reference standard (Table 1A, Figure S1) and considering any LFIA positive result (IgM, IgG or both) as positive. The LFIA devices achieved sensitivity ranging from 55% (95%CI 36-72%) to 70% (51-84%) and specificity from 95% (95%CI 86-99%) to 100% (94-100%). There was no evidence of differences between the devices in sensitivity (p≥0.015, cf. Benjamini-Hochberg p=0.0014 threshold) or specificity (p≥0.19 for all devices with at least one false-positive test). Restricting to 31 samples collected ≥10 days post symptom-onset (all ELISA IgG-positive), LFIA sensitivity ranged from 61% (95%CI 39-80%) to 88% (68-97%) (Table S5).

**Table 1.** Results of nine lateral flow immunoassays (LFIA) devices and an ELISA assay, tested with plasma classified as positive (RT-PCR positive) and negative (pre-pandemic). n=91-182 per LFIA device. Different manufacturers are designated 1-9. 95% confidence intervals (CI) are presented for each point estimate. Any LFIA positive result (IgM, IgG or both) was considered positive. ELISA positive samples were all positive for IgG, no sample was IgM-positive and IgG-negative.

| Assay | RT-PCR positive |  | Pre-pandemic control |  | Sensitivity<br>(95% CI) | Specificity<br>(95% CI) |
| --- | --- | --- | --- | --- | --- | --- |
|  | True | False | True | False |  |  |
|  | positive | negative | negative | positive |  |  |
| ELISA | 34 | 6 | 50 | 0 | 85 (70,94) | 100 (93,100) |
| 1 | 18 | 15 | 60 | 0 | 55 (36,72) | 100 (94,100) |
| 2 | 23 | 15 | 90 | 1 | 61 (43,76) | 99 (94,>99) |
| 3 | 21 | 12 | 58 | 2 | 64 (45,80) | 97 (88,>99) |
| 4 | 25 | 13 | 59 | 1 | 66 (49,80) | 98 (91,>99) |
| 5 | 19 | 12 | 58 | 2 | 61 (42,78) | 97 (91,>99) |
| 6 | 20 | 11 | 59 | 1 | 65 (45,81) | 98 (91,>99) |
| 7 | 23 | 10 | 57 | 3 | 70 (51,84) | 95 (86,>99) |
| 8 | 18 | 14 | 60 | 0 | 56 (38,74) | 100 (94,100) |
| 9 | 22 | 18 | 138 | 4 | 55 (38,74) | 97 (93,>99) |

### Detection of SARS-CoV-2 antibodies by LFIA vs. ELISA

We also considered performance relative to ELISA (Table S6, Figure S1), because the LFIA devices target the same antibodies. We considered patients positive by this alternative standard if their IgG OD reading exceeded the threshold above (since no samples were IgM-positive, IgG-negative). Sensitivity of antibody detection by LFIA ranged from 65% (95%CI 46-80%) to 85% (66-96%) and specificity from 93% (95%CI 83-98%) to 100% (94-100%); however, the device with the highest sensitivity had one of the lowest specificities (Figure S1). There was no evidence of differences in sensitivity (p≥0.010, cf. p=0.0014 threshold) or specificity between devices (p≥0.19).

Of 50 designated negative samples tested by both ELISA and the nine different LFIA devices, nine separate samples generated at least one false-positive, on seven different LFIA devices (Figure 3). Four samples generating false-positive results did so on more than one LFIA device, despite the absence of quantifiable IgM or IgG on ELISA, potentially suggesting a specific attribute of the sample causing a cross-reaction on certain LFIA platforms.

Of the 22 RT-PCR-positive samples collected in the acute setting, six fell below the ELISA detection threshold for IgM or IgG; two of these six were positive on LFIA testing, each on one (different) device. Of the remaining 16 acute samples (all ELISA IgG-positive), only nine were consistently positive across all nine LFIA devices. Due to limited availability of LFIA devices, fewer tests were performed on the 18 convalescent samples with available ELISA data, all with quantifiable IgG (Figure 2B;3A). Two had no antibody detected on any LFIA device, and only eight were consistently positive across all LFIA devices tested (between 1 and 9 devices tested per sample).

## DISCUSSION

We here present the performance characteristics of a novel ELISA and nine selected LFIA devices for detecting SARS-COV-2 IgM and IgG. Among 40 RT-PCR-confirmed positive patients, 85% had IgG detected by ELISA, including 100% patients tested ≥10 days after symptom onset. A panel of LFIA devices had sensitivity between 55 and 70% against the reference-standard RT-PCR, or 65-85% against ELISA, with specificity of 95-100% and 93-100%, respectively. These estimates come with wide confidence intervals due to constraints on the number of devices made available. Comparable results have been obtained through a similar appraisal undertaken independently, in which specificity ranged from 84%-100.0%, and the proportion of specimens testing positive increased over time from symptom onset, with >80% sensitivity achieved by some LFIA devices at later time points [15]. Our study, and these parallel data from another centre [15], provide a benchmark against which to assess the performance of future antibody testing platforms, with the aim of guiding decisions about deploying antibody testing and informing the design of second-generation assays.

LFIA devices are cheap to manufacture, store and distribute, and could be used as a point-of-care test, offering an appealing approach to diagnostics and evaluating exposure. A positive antibody test is currently regarded as a probable surrogate for immunity to reinfection. Secure confirmation of antibody status would therefore reduce anxiety, provide confidence to allow individuals to relax social distancing measures, and guide policy-makers in the staged release of population lock-down, potentially in tandem with digital approaches to contact tracing [16]. As a diagnostic tool, serology may have a role in combination with RT-PCR testing to improve sensitivity, particularly of cases presenting some time after symptom onset [17,18]. Reproducible methods to detect and quantify vaccine-mediated antibodies are also crucial as COVID vaccines enter clinical trials.

Appropriate thresholds for sensitivity and specificity depend on the primary purpose of the test. For diagnosis in symptomatic patients, high sensitivity is required (generally ≥90%). Specificity is less critical as some false-positives could be tolerated (provided other potential diagnoses are considered, and accepting that over-diagnosis causes unnecessary quarantine or hospital admission). However, if antibody tests were deployed as an individual-level approach to inform release from quarantine, then high specificity is essential, as false-positive results return non-immune individuals to risk of exposure. For this reason, the UK Medicines and Healthcare products Regulatory Agency has set a minimum 98% specificity threshold for LFIAs [19].

Appraisal of test performance should also consider the influence of population prevalence, acknowledging that this changes over time, geography and within different population groups. The potential risk of a test providing false reassurance and release from lock-down of non-immune individuals can be considered as the proportion of all positive tests that are wrong. Based on the working ‘ best case’ scenario of a LFIA test with 70% sensitivity and 98% specificity, the proportion of positive tests that are wrong is 35% at 5% population seroprevalence (19 false-positives/1000 tested), 10% at 20% seroprevalence (16 false-positives/1000) and 3% at 50% seroprevalence (10 false-positives/1000) (Figure 4).

More data are needed to investigate antibody-positivity as a correlate of protective immunity. Indeed pre-existing IgG could enhance disease in some situations [20], with animal data demonstrating that SARS-CoV anti-spike IgG contributes to a proinflammatory response associated with lung injury in macaques [21].

Our study, and another undertaken independently in parallel [11], demonstrates accurate performance of ELISA targeting anti-spike protein antibodies. Additionally, our ELISA results are supported by context regarding disease severity and the time of sampling relative to symptom onset. Our data on the kinetics of antibody responses build upon studies of hospitalised patients in China reporting a median 11 days to seroconversion for total antibody, with IgM and IgG seroconversion at days 12 and 14 respectively [17], and others that report 100% IgG positivity by 15-19 days [18] [22]. Our ELISA data show IgG titres rose over the first 3 weeks of infection and that IgM testing identified no additional cases. Methods to enhance sensitivity, especially shortly after symptom onset, could consider different sample types (e.g. saliva), different antibody classes (e.g. IgA) [23], T-cell assays or antigen detection [24]. In contrast to others [18,25-27], we did not find evidence of an association between disease severity and antibody titres. We observed several LFIA false positives, which may have potentially resulted from cross-reactivity of non-specific antibodies (e.g. reflecting past exposure to other seasonal coronavirus infections).

The main study limitation is that numbers tested were too small to provide tight confidence intervals around performance estimates for any specific LFIA device. Expanding testing across diverse populations would increase certainty, but given the broadly comparable performance of different assays, the cost and manpower to test large numbers may not be justifiable. Demonstrating high specificity is particularly challenging; for example, if the true underlying value was 98%, 1000 negative controls would be required to estimate the specificity of an assay to +/-1% with approximately 90% power. Full assessment should also include a range of geographical locations and ethnic groups, children, and those with immunological disease including autoimmune conditions and immunosuppression.

In summary, antibody testing is a crucial component of measures that may be required to inform release from lockdown. Our findings suggest that while current LFIA devices may provide some information for population-level surveys, their performance is inadequate for most individual patient applications. The ELISA we describe is currently being optimised and adapted to run on a high-throughput platform and provides promise for the development of reliable approaches to antibody detection that can support decision making for clinicians, the public health community, policy-makers and industry.

## Data Availability

Results generated for all samples and relevant metadata is provided in Table S7.

https://doi.org/10.12688/wellcomeopenres.15927.1

## DATA AVAILABILITY

Results generated for all samples and relevant metadata is provided in Table S7.

### ACKNOWLEDGEMENTS

This work uses data and samples provided by patients and collected by the NHS as part of their care and support. We are extremely grateful to the frontline NHS clinical and research staff and volunteer medical students, who collected this data in challenging circumstances; and the generosity of the participants and their families for their individual contributions.

## AUTHOR CONTRIBUTIONS

Leadership roles were undertaken by DC (project management), RJP and DJR (management of pre-pandemic negative sample bank), AJM (identification and recruitment of cases), DWE and PCM (oversight of LFIA, data analysis and manuscript writing), GRS (ELISA design, development and data generation). All authors contributed as follows: JIB, JKB, DC, DWE, PCM, AJM, TEAP, GRS, MGS, ASW conceived and designed the study. EA, RA, MIA, MAi, JKB, SB, EC, TdS, AE, DG, AH, KJ, RL, UL, HM, AJM, JMa, JMi, MFM, SM, MO, RJP, AP, EP, TPr, DR, AS, MGS, DSk, JS, TS, VS, HT, PT, LT, CW, NW collected the clinical data and patient samples. KA, SKC, WD, KED, CD, AK, CL, CLC, JMo, JR, DSt, GRS, JSC, PS, BW obtained the laboratory data. DWE, PCM, TEAP, ASW analysed the data. DC, DWE, PCM, TEAP, ASW wrote the manuscript. EB, JB, TB, MC, RC, DC, HF, SH, PK, JK, CK, PCM, AJM, FP, DSt, GRS, ENS, JW provided study management and governance. All authors reviewed and approved the final version of the manuscript for submission.

## AUTHOR LIST AND AFFILIATIONS

In alphabetical order:

Emily Adams^1^, Mark Ainsworth^2^, Rekha Anand^3^, Monique I Andersson^2^, Kathryn Auckland^4^, J Kenneth Baillie^5^, Eleanor Barnes^2^,^4^, Sally Beer^2^, John I Bell^4^, Tamsin Berry^6^, Sagida Bibi^7^, Miles Carroll^4^,^8^, Senthil K Chinnakannan^4^, Elizabeth Clutterbuck^7^, Richard J Cornall^2^,^4^, Derrick W Crook^2^,^4^, Thushan de Silva^9^, Wanwisa Dejnirattisai^4^, Kate E Dingle^4^, Christina Dold^7^, Alexis Espinosa^2^, David W Eyre^2^,^4^, Helen Farmer^6^, Maria Fernandez Mendoza^2^, Dominique Georgiou^2^, Sarah J Hoosdally^4^, Alistair Hunter^10^, Katie Jeffery^2^, Dominic Kelly^7^, Paul Klenerman^2^,^4^, Julian Knight^2^,^4^, Clarice Knowles^6^, Andrew J Kwok^4^, Ullrich Leuschner^11^, Robert Levin^12^, Chang Liu^4^, César López-Camacho^4^, Jose Martinez^2^, Philippa C Matthews^2^,^4^, Hannah McGivern^13^, Alexander J Mentzer^2^,^4^, Jonathan Milton^13^, Juthathip Mongkolsapaya^4^, Shona C Moore^14^, Marta S Oliveira^13^, Fiona Pereira^15^, Elena Perez^2^, Timothy Peto^2^,^4^, Rutger J Ploeg^2^,^13^, Andrew Pollard^2^,^7^, Tessa Prince^14^, David J Roberts^11^, Justine K Rudkin^4^, Veronica Sanchez^2^, Gavin R Screaton^4^, Malcolm G Semple PhD^14^, Jose Slon-Campos^4^, Donal T Skelly^2^,^16^, Elliot Nathan Smith^6^, Alberto Sobrinodiaz^2^, Julie Staves^2^, David I Stuart4,^17^, Piyada Supasa^4^, Tomas Surik^13^, Hannah Thraves^2^, Pat Tsang^11^, Lance Turtle^14^, A Sarah Walker^4^, Beibei Wang^4^, Charlotte Washington^3^, Nicholas Watkins^18^, James Whitehouse6

1 Liverpool School of Tropical Medicine, Liverpool, L3 5QA, UK

2 Oxford University Hospitals NHS Foundation Trust, Oxford, OX3 9DU, UK

3 NHS Blood and Transplant Birmingham, Vincent Drive, B15 2SG, UK

4 Nuffield Department of Medicine and NIHR Oxford Biomedical Research Centre, University
of Oxford, OX3 9DU, UK

5 Roslin Institute, University of Edinburgh, EH25 9RJ, UK

6 Department of Health and Social Care, UK Government, London, UK

7 Department of Paediatrics, Oxford Vaccine Group, University of Oxford, OX3 7LE

8 Porton Down, Public Health England, Salisbury, SP4 0JG

9 Department of Infection, Immunity and Cardiovascular Disease, The Medical School,
University of Sheffield, Sheffield, S10 2RX, UK

10 NHS Blood and Transplant Basildon, Burnt Mills Industrial Estate, Basildon, SS13 1FH,
UK

11 NHS Blood and Transplant Oxford, John Radcliffe Hospital, Oxford, OX3 9DU, UK

12 Worthing Hospital, Worthing, BN11 2DH, UK

13 Nuffield Department of Surgical Sciences, University of Oxford, OX3 9DU, UK

14 NIHR Health Protection Research Unit in Emerging and Zoonotic Infections, Faculty of
Health and Life Sciences, University of Liverpool, Liverpool, UK
13

15 Imperial College, London, SW7 2AZ, UK1

16 Nuffield Department of Clinical Neurosciences, University of Oxford, OX3 9DU, UK

17 Diamond Light Source, Harwell Science and Innovation Campus, OX11 ODE, UK

18 NHS Blood and Transplant Cambridge, Long Road, Cambridge, CB2 0PT, UK

## FUNDING

This study was supported by the National Institute for Health Research (NIHR) Oxford Biomedical Research Centre, the UK Government Department of Health and Social Care and and grants from NIHR [award CO-CIN-01] and the Medical Research Council [grant MC_PC_19059]. DC, TEAP and ASW are supported by the National Institute for Health Research (NIHR) Health Protection Research Unit in Healthcare Associated Infections and Antimicrobial Resistance at the University of Oxford in partnership with Public Health England (NIHR200915). Blood donor and QUOD samples were provided with support from NHS Blood and Transplant and the Medical Research Council UK. SKC is supported by Medical Research Council UK. TdS is funded by a Wellcome Trust Intermediate Clinical Fellowship (110058/Z/15/Z). DWE is a Robertson Foundation Fellow and NIHR Oxford BRC Senior Research Fellow. PCM is a Wellcome Trust Clinical Research Fellow (110110/Z/15/Z) and NIHR Oxford BRC Senior Research Fellow. EA, AM, PK, SCM, TPr, MGS and LT are supported by NIHR Health Protection Research Unit in Emerging and Zoonotic Infections (HPRU-EZI) at University of Liverpool in partnership with Public Health England (PHE), in collaboration with the University of Oxford and Liverpool School of Tropical Medicine (award number NIHR200907). EB, PK, AJP and ASW are NIHR Senior Investigators. PK (WT109965/MA) and GRS (095541/A/11/Z) are Wellcome Trust Senior Investigators. JR is supported by a Sir Henry Dale Fellowship, jointly funded by the Wellcome Trust and the Royal Society (Grant 101237/Z/13/B). The views expressed are the author(s) and are not necessarily those of the NHS, the NIHR, the UK Department of Health and Social Care, the MRC or PHE.

## COMPETING INTERESTS

RC reports personal fees and other from MIROBIO Ltd, outside the submitted work. DWE reports personal fees from Gilead, outside the submitted work. SH reports grants from NIHR, during the conduct of the study. AJP reports grants from NIHR Oxford Biomedical Research Centre, outside the submitted work; and AJP is Chair of UK Dept. Health and Social Care’ s (DHSC) Joint Committee on Vaccination & Immunisation (JCVI) and is a member of the WHO’ s SAGE. The views expressed in this article do not necessarily represent the views of DHSC, JCVI, NIHR or WHO. GRS reports personal fees from GSK Vaccines SAB. MGS reports grants from National Institute of Health Research, grants from Medical Research Council UK, grants from Health Protection Research Unit in Emerging & Zoonotic Infections, University of Liverpool, during the conduct of the study; other from Integrum Scientific LLC, Greensboro, NC, USA, outside the submitted work. ASW reports grants from NIHR, during the conduct of the study. No other author has a conflict of interest to declare.

## FIGURE LEGENDS

**Figure 1. Cartoon to illustrate the generation of IgM and IgG antibodies to SARS nCoV-2 and detection of antibodies by a lateral flow device**. (A) In vivo generation of antibodies to the trimeric SARS-CoV-2 spike protein. (B) Projected change in titres of specific IgM and IgG over time following infection, with arrows indicating typical time frames for collection of acute and convalescent samples. (C) Ex vivo detection of IgG and/or IgM using a lateral flow immunoassay (LFIA): S= sample well, T=test antibody; C=control. Diagram shows a positive sample on the left, with positive lines at both C and T, and a negative test on the right with a line present only at C. Any other combination of lines renders the test invalid. Some devices have two test lines, for separate detection of anti-SARS-nCoV-2-IgG and -IgM. (D) Outcomes of testing negative and positive plasma using LFIA. (E) Calculation of sensitivity, specificity, positive and negative predictive value of a test. Image created with BioRender.com; exported under a paid subscription.

**Figure 2: Results of testing 90 plasma samples for SARS-CoV-2 IgM and IgG by Enzyme linked Immunosorbent Assay (ELISA)**. (A) IgM readings for SARS-CoV-2 pre-pandemic plasma (designated negatives, shown in blue, n=50), and RT-PCR confirmed cases of SARS-CoV-2 infection (designated positives, shown in orange, n=40; divided into acute cases, n=22, and convalescent cases, n=18. Threshold of OD = 0.07 discriminates accurately between negative controls and convalescent sera. (B) IgG data shown for the same subgroups described for panel A. A threshold of OD = 0.4 discriminates between designated negatives and positives. (C) IgM OD values plotted against the time post symptoms at which plasma was obtained. The line shows the mean OD value expected from a spline-based linear regression model, the ribbon indicates the pointwise 95% confidence interval. (D) IgG OD values plotted against the time post symptoms at which plasma was obtained. Coloured dots in panels C and D indicate disease severity. OD = optical density.

**Figure 3: Comparison between ELISA and LFIA for SARS-CoV-2 designated negative and positive plasma**. Panel A shows quantitative optical density (OD) readout from ELISA for IgG for designated negative plasma (n=50) and from individuals with RT-PCR confirmed infection (n=40, divided into acute and convalescent plasma). IgM results are shown in Figure S2 in the Supplementary Materials. Panel B shows results from LFIA produced by nine manufacturers. Any positive test for IgG, IgM, both or total antibody is shown as positive, please see Figure S2 for more detailed breakdown. Grey blocks indicate missing data as a result of insufficient devices to test all samples and one assay on one device with an invalid result. Samples in both panels are ranked from left to right by quantitation of IgG (as indicated in panel A).

**Figure 4: Influence of population prevalence of seropositivity on assay performance**. Scenarios with population prevalence of 5%, 20% and 50% are shown within each panel. Panel A shows the proportion of all positive tests that are wrong (1-positive predictive value), which would lead to false release from lock-down of non-immune individuals, for varying test sensitivity (x-axis) and 1-specificity (line colour). Panel B shows the proportion of negative tests that are wrong, panel C the absolute number of false positive tests per 1000 tests and panel D the absolute number of false negative tests per 1000 tests.

## SUPPLEMENTARY MATERIAL

**Supplementary Figure S1: Sensitivity and specificity of lateral flow devices compared with RT-PCR confirmed cases and pre-pandemic controls (panels A and B) and compared with ELISA results (panels C and D)**.

**Supplementary Figure S2: Comparison between ELISA and LFIA for SARS-CoV-2 designated negative and positive plasma**.

**Supplementary table S1. STARD checklist** (provided as a separate pdf)

**Supplementary table S2. Metadata describing origin and characteristics of designated negative controls and individuals with confirmed SARS-CoV-2 infection** (provided as a separate .xlsx file)

**Supplementary table S3. Summary grid presenting the number of samples from each cohort tested using different assay platforms**.

**Supplementary table S4. Multivariable regression models for relationship between ELISA IgM and IgG readings and covariates in RT-PCR positive cases**.

**Supplementary Table S5. Results of nine lateral flow immunoassays (LFIA) devices and an ELISA assay, tested with plasma classified as positive (RT-PCR positive) obtained from patients** ≥**10 days after onset of symptoms**.

**Supplementary Table S6. Results of nine lateral flow immunoassays (LFIA) devices, tested with plasma classified as positive and negative using ELISA as an alternative reference standard (n=81-90 per LFIA device)**. Different manufacturers are designated A-95% confidence intervals (CI) are presented for each point estimate.

**Supplementary table S7: Results of all assays performed and relevant metadata**. (provided as a separate .xlsx file)

## Notes

### Competing Interest Statement

None

